# The whole day matters after stroke: Study protocol for a randomized controlled trial investigating the effect of a ‘sit less, move more, sleep better’ program early after stroke

**DOI:** 10.1101/2023.08.11.23293976

**Authors:** Deborah Okusanya, Joy Ezeugwa, Aiza Khan, Brian Buck, Glen Jickling, Victor Ezeugwu

## Abstract

**Background:** Movement-related behaviours, including prolonged sedentary behaviour, physical inactivity, and sleep disorders, are associated with worse functional outcomes poststroke. Addressing these co-dependent behaviours early after stroke may help to optimize recovery and improve overall quality of life for individuals with stroke.

**Objective:** This study aims to determine the feasibility and effect of a ‘sit less, move more, sleep better’ program early after stroke on functional mobility and global disability outcomes, while also exploring imaging and behavioral markers that may influence walking recovery.

**Methods:** The study is a single-blind (outcome assessor), single-center, parallel-group, randomized controlled trial. We will enroll 50 patients with acute ischemic stroke within 7 days from symptom onset, aged 18 years or older, and with ongoing walking goals. Demographic and stroke characteristics, including stroke risk factors, neuroimaging, and acute stroke treatments, will be determined and documented. All participants will wear an accelerometer for one week at three different time-points (baseline, 6, and 12 weeks) to assess movement-related behaviours.

Following randomization, participants in the intervention arm will receive a ‘sit less, move more, sleep better’ program for up to 1 hour/day, 5 days/week, for 6 weeks to enhance self-efficacy for change. Participants in the control arm will receive usual inpatient and early supported stroke discharge care. The feasibility outcomes will include reach (enrolled/eligible), retention (completed/enrolled), adverse events, and program adherence. Other outcomes at 6 and 12 weeks include the modified Rankin Scale, Timed-Up and Go, movement-related behaviours, walking endurance, gait speed, cognition, stroke severity and quality of life.

**Discussion:** Adopting a whole-day approach to poststroke rehabilitation will provide valuable insights into the relationship between optimizing movement-related behaviours early after stroke and their impact on functional outcomes. Through exploring person-specific behavioural and imaging markers, this study may inform precision rehabilitation strategies, and guide clinical decision making for more tailored interventions.

## Background and rationale

The main aim of rehabilitation is to enhance recovery trajectories and improve adaptation to the loss of neurologic function, with the goal of promoting function and preventing stroke recurrence [1]. Stroke is a heterogeneous disorder characterized by diverse brain and behavioural changes, as well as varied rehabilitation outcomes among survivors. This heterogeneity presents a significant challenge when attempting to employ a “one-size-fits-all” approach to stroke rehabilitation [2]. Integrating knowledge about mechanisms of stroke recovery, biomarkers, and motor outcomes is a key challenge to inform individualized care and the development of targeted interventions.

Motor recovery begins early after stroke through neuroplasticity, with the greatest improvements occurring within the first 3 to 6 months after injury [3,4]. Therefore, optimizing rehabilitation during this critical window of recovery is important to enhance the recovery process. Preclinical animal studies have shown that more task-specific reaching movement repetitions are associated with better functional recovery after stroke, with upwards of 600-700 repetitions per day suggested in models with the most severe impairments [5]. However, the optimal dosage of stepping repetitions for humans is still undetermined. Moreover, people with stroke are largely sedentary throughout their acute and inpatient rehabilitation journey [6,7]. In our previous study, we showed that people with stroke, following inpatient rehabilitation and transitioning back to the community, spend approximately three-quarters of their waking hours in sedentary behaviour, and half of the participants have long sleep durations exceeding 9 hours per day [8]. Subsequently, we demonstrated that it is feasible to enhance self-efficacy and decrease sedentary time after stroke [9].

To inform personalized stroke rehabilitation, accelerometry may be utilized in addition to clinical outcomes to understand daily movement-related behaviours (sleep, sedentary behaviour, and physical activity) that influence stroke recovery. The use of wearable activity sensors enables unsupervised, continuous, and objective monitoring of performance during and outside structured therapy [10]. By recording exercise and non-exercise tasks over the whole day, a representative and reliable account of performance is obtained with minimal therapist burden. Studying whole-day movement-related behaviours can provide valuable insights into the complex interplay of the 24-Hour Activity Cycle [11]. Clearly, increasing time spent in one behaviour displaces time in the other behaviours within a 24-hour period [12]. For instance, dedicating more time to physical activity necessitates reducing time spent sleeping or engaging in sedentary behaviour, or both. Gaining insight into these relationships is crucial for targeted care, as they are predictors of recovery and health outcomes [13,14]. Employing a whole-day approach that maximizes stepping time and repetitions, frequently interrupts sedentary behaviour, and optimizes sleep may be beneficial.

In addition to behavioural outcomes, an understanding of novel imaging biomarkers may also guide intervention development for personalized stroke rehabilitation. In addition to cortical infarct volume, leukoaraiosis or cerebral white matter disease, recognizable on magnetic resonance imaging (MRI) as areas of hyperintensities [15] has gained significance as a potential moderator of stroke recovery [16–18]. In older adults, leukoaraiosis is associated with impairments in gait parameters, including step length and cadence, possibly due to ongoing axonal changes [19,20]. After a stroke, the impact of leukoaraiosis becomes more evident [21], possibly due to a diminished capacity of the cerebral tissue to respond to the brain injury [22]. For example, following mechanical thrombectomy poststroke, only 24% of patients with severe leukoaraiosis had a “good” 90-day modified Rankin Scale (mRS) score compared to 85% of patients without leukoaraiosis [18]. To date, there are no known therapies to slow the progression of leukoaraiosis and no rehabilitation interventions have been specifically developed for survivors of stroke with leukoaraiosis. Understanding the role of leukoaraiosis as a moderator of stroke recovery may help to identify individuals that may benefit from a different approach to rehabilitation through the whole-day strategy. We hypothesize that a whole-day rehabilitation approach will optimize movement-related behaviours and improve functional mobility after stroke, moderated by the extent of cortical infarct volume and leukoaraiosis.

## Methods

### Study design and setting

The study is a single-blind (outcome assessor), single-center, parallel-group, randomized controlled trial. The Standard Protocol Items: Recommendations for Interventional Trials (SPIRIT) guidelines will be followed for this study [23]. The study has received ethics approval from the Human Research Ethics Board at the University of Alberta (Pro00127990, February 3, 2023) and the protocol has been registered on the clinical trial registry (www.ClinicalTrials.gov ID: NCT05753761, March 3, 2023). Written informed consent will be obtained from the potential participant. The schedule of enrolment, interventions, and assessments is shown in **Fig 1**. The study will be conducted at the University of Alberta Hospital in Edmonton, Alberta, Canada and is anticipated to be completed within 24 months beginning from July 12, 2023 to June 30, 2025.

**Fig 1.**
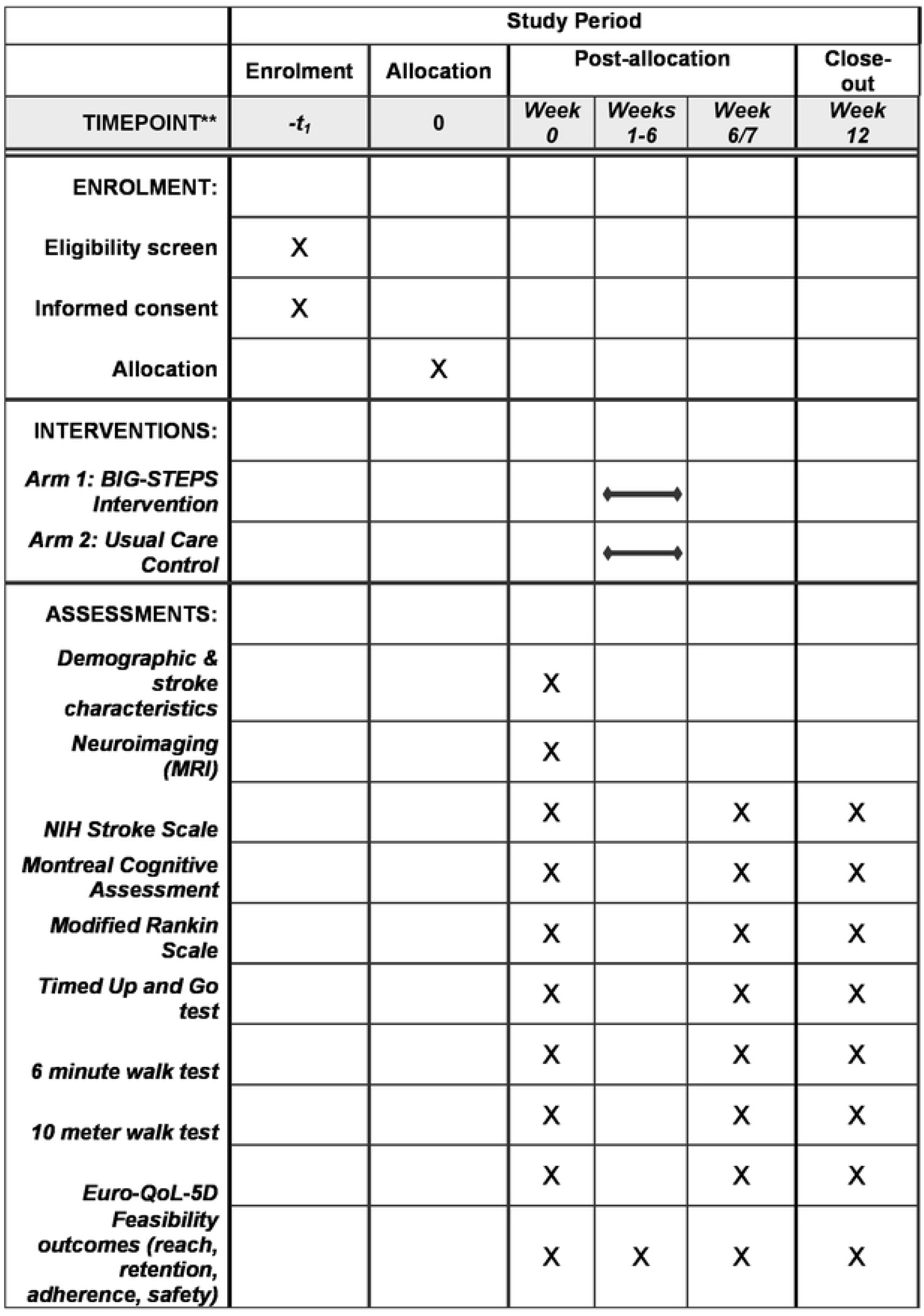
Study schedule. BIG STEPS, Behaviour and Imaging Guided Stepping Training Early Post-Stroke.

### Patient population and eligibility criteria

Eligible participants will be within 7 days of ischemic stroke onset, aged 18 years or older, deemed medically stable by physicians, and able to walk at least 5 meters with or without a gait aid and minimal assistance from 1-person, and able to understand and follow instructions. Additionally, eligible participants will have ongoing mobility deficits such as foot drop, knee buckling, or gait asymmetry or other walking challenges such as walking speed below 1.0 meters per second. Exclusion criteria include previously diagnosed mobility-limiting conditions such as multiple sclerosis or Parkinson’s disease, or if they have active cancer, uncontrolled high blood pressure, or an unstable cardiovascular condition. Rehabilitation team leads and care management staff will assist in study recruitment by distributing consent to contact forms to potential participants. After receiving informed consent, information regarding demographics, stroke characteristics, risk factors, infarct location and volume, and acute stroke treatments received (e.g. thrombectomy) will be determined and documented. Subsequently, participants will be set up to wear an activPAL accelerometer for one week, validated in stroke and that accurately distinguishes sedentary from non-sedentary behaviours [24]. The activPAL will be worn on the less affected thigh, separated from the skin by a soft cotton pad, and secured with a transparent film dressing to follow a 24-hour protocol.

## Interventions

### Experimental Group: Behaviour and Imaging-Guided Stepping Training Early Post-Stroke (BIG STEPS)

In addition to standard of care management for ischemic stroke according to the Canadian Stroke Best Practices Guidelines [25], participants allocated to the experimental group will receive a ‘sit less, move more, sleep better’ intervention focused on reducing and frequently interrupting sedentary time, taking more steps, and sleeping better. Data acquired from the baseline activPAL monitoring for 7 days will be discussed with the participants and used to inform personalized whole-day activity and behaviour-change recommendations, including replacing prolonged sedentary periods with stepping time. **Fig 2** shows examples of outputs from the activPAL monitor before and after a behaviour change intervention focused on reducing and interrupting sedentary behaviour throughout the day. Guided by the Social Cognitive Theory framework [26], behaviour-change coaching sessions, outside of structured therapy, will be conducted in person or virtually (where applicable), for up to 1 hour a day, 5 days a week for 6 weeks to enhance self-efficacy to ‘sit less, move more, and sleep better.’ Each week, the stepping goal will be increased according to the previous week’s activity level and the participant’s baseline activity. For instance, if the participant averaged 1000 steps daily at baseline, the focus of the coaching and recommended strategies will be to increase steps to 1500 steps daily over the next week, depending on the participant’s ability. Based on the participant’s progress, the stepping goal will be incrementally increased up to 6000 steps per day over 6 weeks. Following the Canadian Movement Guidelines of 7-9 hours of good quality sleep [27], sleep time will also be discussed and strategies to optimize sleep included in the goals for each week, such as establishing a routine bedtime schedule or avoiding screen-based activities about 30-60 minutes before bedtime.

**Fig 2.**
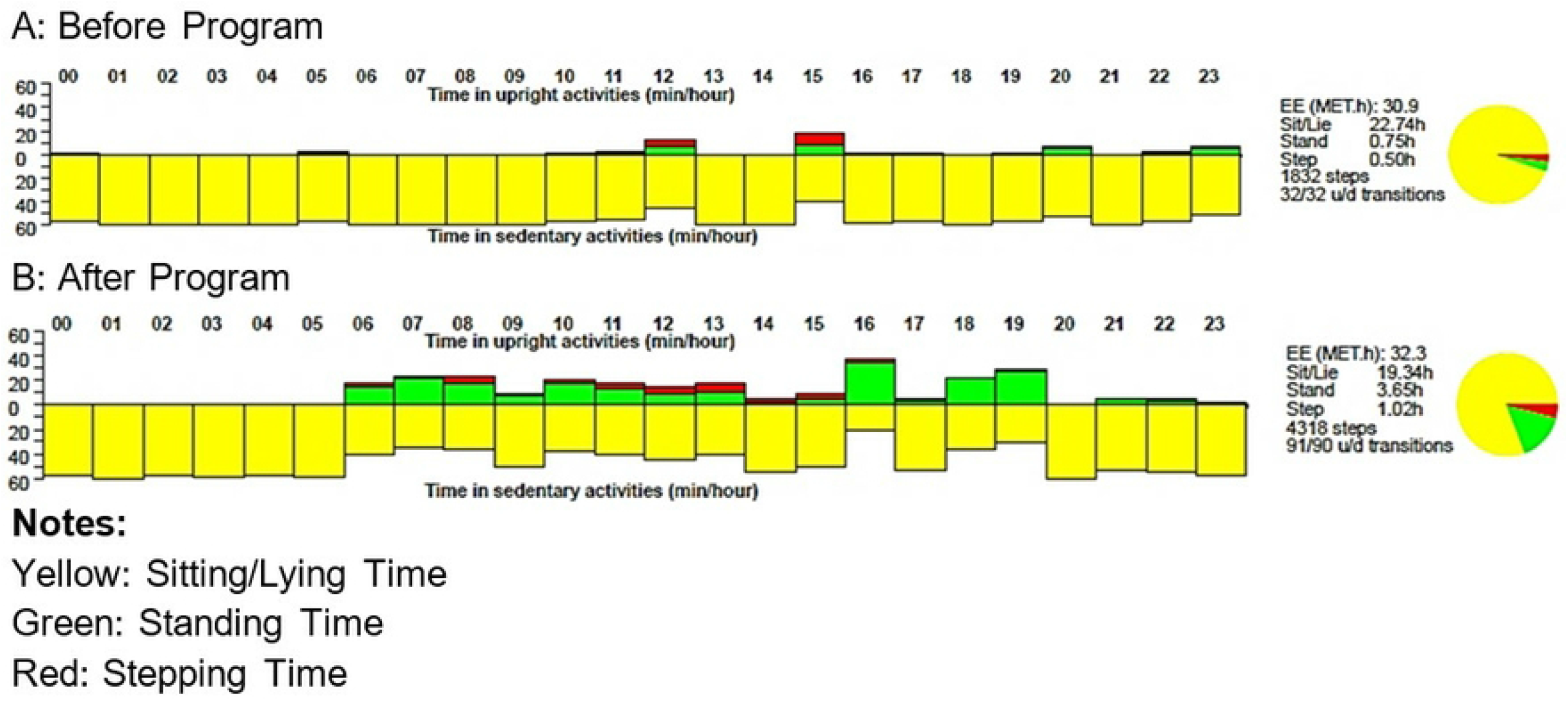
Examples of movement-related behaviours before and after a behaviour change program.

Participants in the BIG-STEPS arm will also wear a step counter on their ankle (Fitbit Inspire, Alphabet Inc., USA) [28] for self-monitoring, daily feedback and to monitor adherence. If a participant is discharged from the hospital before 6 weeks, we will complete the follow-up visits at the discharge location or virtually (where applicable).

### Control Group: Usual Care Intervention

Participants allocated to the control group will receive usual inpatient and early supported discharge care (for those discharged early from hospital) in accordance with the Canadian Stroke Best Practices Guidelines [25]. This can include therapeutic mobilization directed by physical therapists or general mobilization directed by nursing staff, as tolerated. The specific exercises and treatments chosen for therapy will be up to the discretion of the therapy team. Study staff will document details regarding therapy sessions and any related issues. Participants in the usual care group will complete all assessments, including activity monitoring with the activPAL at the same time points as the experimental group.

### Randomization

Prior to the intervention, participants will be randomized on a 1:1 ratio, stratified by age (<60 or ≥60 years) and sex (male or female) to either the intervention or usual care group using computer-generated permuted random block sizes of 2 or 4 on a web-based platform (studyrandomizer.com).

### Blinding

Allocation will be fully concealed and completed by a blinded study team member not involved in data collection. Outcomes assessors will be blinded to group allocation.

## Data Collection and Management

Participant demographic, vital signs, and clinical characteristics, including stroke risk factors, infarct location and volume, and acute stroke treatments received (e.g. thrombectomy) will be determined and documented confidentially using the Research Electronic Data Capture (REDCap) register. Access to the register will be limited to authorized personnel. Data collection is currently ongoing.

### Outcomes

Feasibility outcomes will be assessed including safety and adverse events, reach, retention/attrition, and adherence/intervention fidelity as outlined in Table 1. Other outcomes will include global disability assessed using the 6-item modified Rankin Scale (mRS) [29] and functional mobility assessed using the Timed Up and Go (TUG) test [30]. Time spent in movement-related behaviours (sleep, sedentary behaviour, and physical activity) will be measured with activPAL accelerometer, gait speed with 10-Meter Walk Test [31], walking endurance with 6-Minute Walk Test [32]. The National Institute of Health Stroke Scale (NIHSS) [33] will be used to classify stroke severity, and the Montreal Cognitive Assessment (MOCA) [34 to assess cognitive impairment. Quality of life will be assessed using the Euro-QOL 5D (EQ-5D) [35].

**Table 1.**
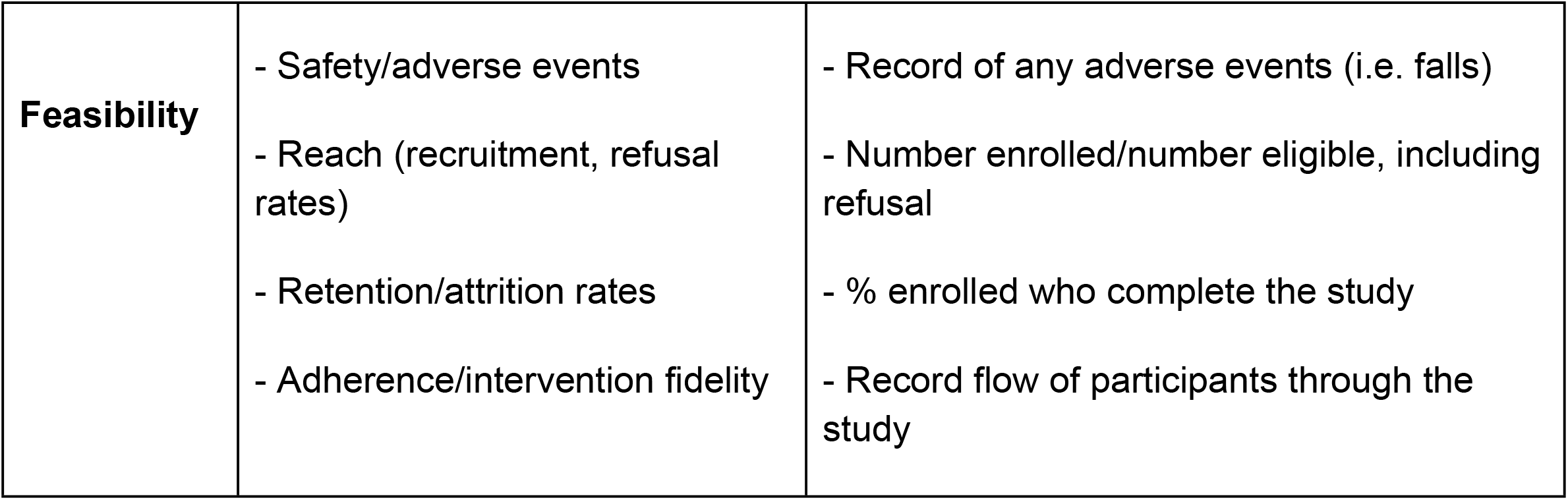
Feasibility metrics.

MRI imaging will be collected only at baseline. Cortical infarct volume and leukoaraiosis will be determined from MRI scans, analyzed using standardized automated segmentation protocol of fluid-attenuated inversion recovery sequences, independent of demographic or clinical data.

### Sample size estimates

The sample size was estimated based on data from a previous feasibility study using an assumed difference of 3.8 seconds on the TUG test [9]. Thus, a sample of 45 participants will achieve a power of 0.81 in repeated measures using GEE at α=0.05. With an anticipated 10% attrition, 50 participants will be recruited.

### Statistical Analysis

Participant baseline characteristics will be compared using independent t tests (continuous variables) or Chi-Square test (categorical variables). Feasibility measurements (safety, reach, retention, adherence) will be calculated. Changes in functional mobility and global disability, movement-related behaviours, functional and quality of life outcomes across time (baseline to immediately post intervention to follow-up) will be determined using mixed-effects models. Compositional associations between movement-related behaviours and functional outcomes will be analyzed using compositional data analysis. We will consider the moderating effect of cortical infarct volume and leukoaraiosis on outcomes.

### Safety and adverse monitoring

Safety concerns were first considered in the feasibility study, in which no adverse events were reported. Patients will be closely monitored by the study team during assessments. Furthermore, before participants complete any walking tests such as the 6-min walk test, a safety screen will be conducted to ensure there are no “red flags” or contraindications to activity. If a patient’s condition worsens significantly or if the patient requests to end their involvement in the study, all data collection will be stopped immediately, and their participation in the study will be terminated.

### Discussion and summary

Despite rapid advances in acute stroke care, stroke remains the leading cause of adult disability globally [36]. It is concerning that survivors of stroke spend most of their time in sedentary behaviours [8], including during stroke rehabilitation [7]. Responsible factors may include poor patient compliance with assigned tasks outside of structured therapy or limited dose of rehabilitation after stroke. A meta-analysis showed that each additional hour of sedentary time, beyond 11 hours per day, increases the risk of stroke by 21% in the general population [37]. The influence of sedentary behaviour on stroke recurrence is unknown. The field of stroke recovery, rehabilitation, and restorative neuroscience is still young, and this project will contribute to our understanding of the effect of a whole-day approach to rehabilitation early poststroke.

Integrating the knowledge around mechanisms of stroke recovery, biomarkers, and development of targeted interventions to inform precision rehabilitation is a main challenge in rehabilitation science. Rehabilitation intervention studies, accounting for the heterogeneity in stroke, will ensure that the right patient receives the right intervention at the right time point after stroke with the right outcome measurement that is responsive to change [38]. Using a combination of behavioural and imaging markers may help to better understand and personalize rehabilitation early after stroke to optimize function and recovery.

The whole-day approach to promote activity, reduce sedentary behaviour, and optimize sleep may be beneficial for many people with stroke, particularly poor responders to conventional rehabilitation poststroke. Personalized intervention using theory-based behaviour change techniques could lead to a paradigm shift in how rehabilitation care is delivered early after stroke. Future research would explore a larger implementation trial of this approach to poststroke rehabilitation.

## Data Availability

No datasets were generated or analyzed during the current study.

## Supporting Information

**S1 Appendix. SPIRIT 2013 checklist**. (Doc)

**S2 Appendix. Study protocol**. (PDF)

## Author Contributions

**Conceptualization**: Brian Buck, Glen Jickling, Victor Ezeugwu

**Data curation:** Joy Ezeugwa, Aiza Khan, Victor Ezeugwu

**Formal analysis:** Victor Ezeugwu

**Funding acquisition:** Victor Ezeugwu

**Investigation:** Deborah Okusanya, Joy Ezeugwa, Aiza Khan, Brian Buck, Glen Jickling, Victor Ezeugwu

**Methodology:** Deborah Okusanya, Joy Ezeugwa, Aiza Khan, Brian Buck, Glen Jickling, Victor Ezeugwu

**Project administration:** Deborah Okusanya, Joy Ezeugwa, Aiza Khan, Brian Buck, Glen Jickling, Victor Ezeugwu

**Resources:** Deborah Okusanya, Joy Ezeugwa, Aiza Khan, Brian Buck, Glen Jickling, Victor Ezeugwu

**Software:** Joy Ezeugwa, Aiza Khan, Victor Ezeugwu

**Supervision:** Brian Buck, Glen Jickling, Victor Ezeugwu

**Validation:** Brian Buck, Glen Jickling, Victor Ezeugwu

**Visualization:** Deborah Okusanya, Victor Ezeugwu

**Writing – original draft preparation:** Deborah Okusanya, Victor Ezeugwu

**Writing – review & editing:** Deborah Okusanya, Joy Ezeugwa, Aiza Khan, Brian Buck, Glen Jickling, Victor Ezeugwu

## Notes

### Competing Interest Statement

The authors have declared no competing interest.

### Clinical Trial

NCT05753761

### Funding Statement

The author(s) received no specific funding for this work.

### Author Declarations

Human Research Ethics Board at the University of Alberta (Pro00127990)

